# *HLA-A*\**02:01* allele is associated with decreased risk and a longer survival in pancreatic cancer: Results from an exhaustive analysis of the *HLA* variation in PDAC

**DOI:** 10.1101/2024.08.29.24312704

**Authors:** Alberto Langtry, Raul Rabadan, Lola Alonso, Casper van Eijck, Teresa Macarulla, Rita T Lawlor, Alfredo Carrato, Rafael Alvarez-Gallego, Mar Iglesias, Xavier Molero, J Matthias Löhr, Christopher W Michalski, José Perea, Michael O’Rorke, Víctor M Barberà, Adonina Tardón, Antoni Farré, Luís Muñoz-Bellvís, Tatjana Crnogorac-Jurcevic, Enrique Domínguez-Muñoz, Thomas Gress, William Greenhalf, Linda Sharp, Sergio Sabroso-Lasa, Ioan Filip, Gaby Strijk, Florian Castet, Joaquim Balsells, Eithne Costello, Jörg Kleeff, Bo Kong, Josefina Mora, Damian O’Driscoll, Aldo Scarpa, Weimin Ye, Francisco X. Real, Núria Malats, Evangelina López de Maturana, PanGenEU Investigators

## Abstract

Genetic susceptibility loci are associated with PDAC risk and survival, but the impact of germline HLA region variation remains largely unexplored. This study examined *HLA* I-II alleles within the PanGenEU study and validated our findings using external datasets (UK Biobank, TCGA, PAN-NGS trial, and Caris trial). *HLA-A*02:01and HLA-B*49* alleles were linked to a decreased risk of PDAC, whereas *HLA-B*39*, *HLA-DPB1*04,* and *HLA-A*26:01* were directly associated with increased risk. PDAC patients carrying the *HLA-A*02:01* allele also showed lower mortality rates, with the effect being more pronounced in those with *KRAS*^G12V^ mutations, pointing to a host*tumor genetic interaction. This research highlights *HLA-A*02:01*, found in 20% of Europeans, as a marker for reduced PDAC risk and mortality, especially in *KRAS*^G12V^ mutated tumors. Results from this study could enhance personalized medicine for PDAC by identifying patients who may benefit from regular screenings through tailored risk assessments. Importantly, our findings are crucial for stratifying PDAC patients based on their genetic background and tumor mutational profile, which can guide treatment strategies.

## INTRODUCTION

Pancreatic cancer has a dismal prognosis with a 5-year survival rate of 5-10% (1–3) and a 10-year survival rate of 1% (4). Worryingly, it is expected to become the second cause of cancer-related deaths in the USA by 2030 if action is not taken (5). Pancreatic ductal adenocarcinoma (PDAC) is the most common form of these neoplasms. So far, great efforts are being made to advance its diagnosis and detect tumors at early stages to improve survival (6–8). Identifying the genetic and non-genetic factors involved in PDAC etiology is crucial for defining high-risk individuals who would benefit from detecting the tumor when it is still curable. However, there is no definition of high-risk individuals for sporadic PDAC, those representing 90% of the burden of the disease.

We previously reported an inverse association between PDAC risk and nasal allergies, asthma, and autoimmune diseases (9,10), conditions characterized by an overactive immune response. Furthermore, our recent multilayered GWAS approach (11) identified a hotspot in the Major Histocompatibility Complex (*MHC*) region, where the alternative allele of the top targeted Human Leukocyte Antigen *(HLA) G* gene was associated with increased PDAC risk (p-value=1.8×10^−4^). In addition, several GWAS hits at this region have been associated with the risk of other cancer types like breast (12) or lung cancer (13,14), as well as with well-established PDAC risk and protective factors such as chronic immune-related diseases (15,16), diabetes (17,18), smoking (19), asthma (20,21), and nasal allergies (22). The lack of GWAS-reported PDAC genetic susceptibility loci in the *MHC* region might be because SNPs are not fully capturing the variability of the *MHC*. On the other hand, at the tumor level, Filip et al. (2023) (23) reported that Allele-Specific Expression loss of the *HLA* class I genes in PDAC, among other cancers, is associated with a worse prognosis.

The *MHC* is essential for the proper response of the immune system against tumor cells (24). The *MHC* genetic complex is a dense cluster of genes located at the 6p21.1-21.3 chromosome cytoband (28,477,797-33,448,354 bp, GRCh37), and it includes three *HLA* sub-regions. *HLA* class I can be classified into classical (*HLA*-A, -B, and -C) and non-classical genes (*HLA*-*E*, -*F*, and -*G*). *HLA* class II genes are *HLA*-*DR*, -*DQ*, and -*DP*. HLA class I and II proteins present intracellular and extracellular peptides to CD8^+^ and CD4^+^ T-cells, respectively (25,26). The *MHC* region also shows an extensive linkage disequilibrium (27). As a consequence of its complexity, the role of this region has not yet been studied in-depth in relation to the risk of cancer and, more specifically, PDAC.

Here, we report an exhaustive study on PDAC genetic susceptibility associated with the *MHC* region variation on *HLA* class I and II alleles, haplotypes, and amino acids (**Fig. 1**). We performed association analyses at the *HLA* allele and haplotype levels in PanGenEU, one of the largest PDAC case-control studies with extensive standardized clinical and epidemiological data annotation. We then tested these associations in an independent population, the UK Biobank (UKB), and computed global risk estimates by meta-analyzing the results from the two consortia. We also deliver for the first time an estimate of the heritability for PDAC risk explained by the *HLA* class I and II variation. Furthermore, we used the resources from TCGA, UKB, and PanGenEU to conduct a meta-analysis on the HLA alleles’ overall survival (OS) associations. Finally, we assessed the association between the most promising alleles and common *KRAS* mutations using TCGA data and validated results with PAN-NGS and Caris trials. Altogether, our findings underscore the role of the *HLA* region in PDAC genetic susceptibility and prognosis. Our findings may have implications for the design of mutant KRAS-based vaccines in PDAC.

**Figure 1.**
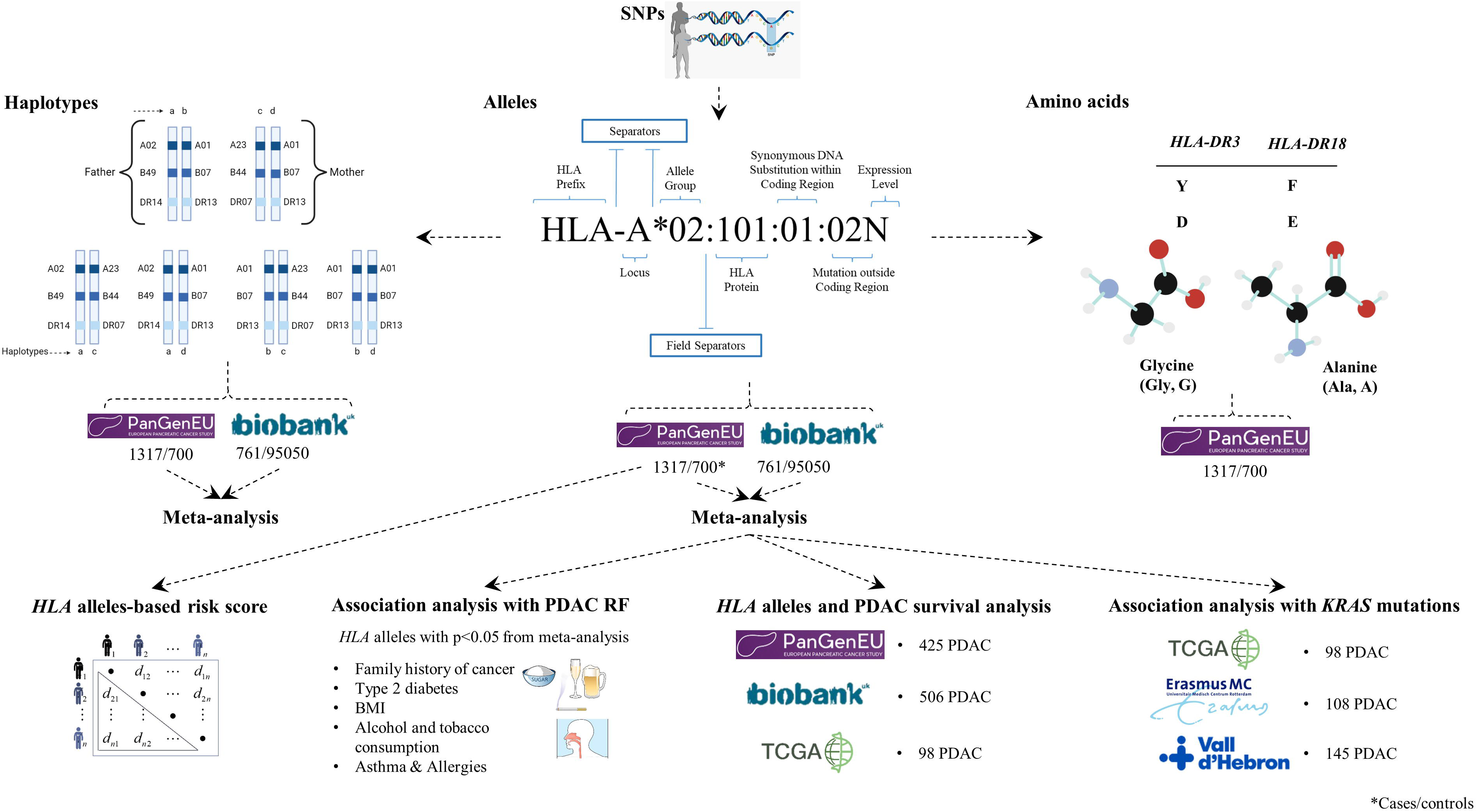
Study Flowchart: Overview of the complementary approaches adopted in this study to explore the role of *MHC* region in the PDAC susceptibility and prognosis.

## RESULTS

### Characteristics of the study populations

The characteristics of the PanGenEU, UK Biobank, and TCGA study populations are available in **Supplementary Table S1**. PanGenEU participants were Caucasian from Spain (58.8%), Italy (22.6%), Sweden (7.9%), Germany (6.7%), UK (3.7%), and Ireland (0.2%). The average age was similar between cases and controls in each study population; however, UKB controls were, on average, younger than cases. A slightly lower proportion of females across the studies was observed for both cases and controls. In the PanGenEU and UKB populations, cases were more frequently smokers and diabetic, while controls had a higher prevalence of asthma and allergies. Finally, the median survival was shorter for UKB patients than for TCGA and PanGenEU patients. As expected based on study design, TCGA included a higher proportion of patients with early-stage tumors than PanGenEU cases. We also observed significant differences in the stage of the tumors across populations, being the Caris clinical trial the population with the highest proportion of stage III-IV PDAC tumors. The median survival time of the UKB patients was significantly shorter compared to the PanGenEU, TCGA, Erasmus, and Caris patients. Finally, neoadjuvant and adjuvant settings were significantly different between the Erasmus and Caris populations.

### Association analyses between HLA alleles and PDAC risk

HLA class I and class II alleles explain 11% of the PDAC risk. Bayesian multi kernel-based regression allowed us to obtain the genetic and residual variance components 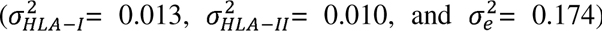 and therefore, the heritability estimates for *HLA* class I and class II alleles were: 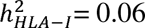 and 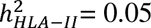.

**Supplementary Fig. S1A** and **S1B** show the landscape of *HLA* class I and II alleles at two-field level in PanGenEU and UKB populations, respectively. *HLA-B* was the most polymorphic *MHC* gene in both populations: 97/298 (32.6%), two-field *HLA* alleles in the PanGenEU and 108/306 (35.3%) in UKB. Some alleles, like *HLA-DPA1*01:03*, showed large variability across populations. In general, the AF matched expected European frequencies (28). The AF in PanGenEU showed good correlations with the T1DGC reference panel (R=0.92 and R=0.86, for one-(allele group) and two-fields (HLA protein) *HLA* alleles, see, **Supplementary Fig. S2A and S2B**) and moderate correlation with UKB (R=0.57, see **Supplementary Fig. S3**). **Supplementary Table S2** summarizes the number of *HLA* class I and II alleles considered in the association analyses after filtering out those with poor imputation quality and low AF.

In our comprehensive meta-analysis of the PanGenEU and UKB populations, the following four alleles yielded significant associations with PDAC risk: *HLA-A*\*02:01 (OR=0.86, p-value=1.88×10^−3^), *HLA-B*\*49 (OR=0.68, p-value=3.81×10^−2^), *HLA-B*\*39 (OR=1.42, p-value=1.99×10^−2^), and *HLA-DPB1*\*04 (OR=1.1, p-value=2.86×10^−2^) (**Fig. 2)**. After multiple testing correction, only the *HLA-A*\*02:01 remained significant (OR=0.86, corrected p-value=2.25×10^−2^, **Supplementary Table S3**). We also explored the association between the above-mentioned *HLA* alleles and the established PDAC risk factors. Interestingly, we observed a significant inverse association between *HLA-B*\*39 and nasal allergies, which is protective against PDAC (OR=0.84, p-value=1.15×10^−4^, see **Fig. 2C**).

**Figure 2.**
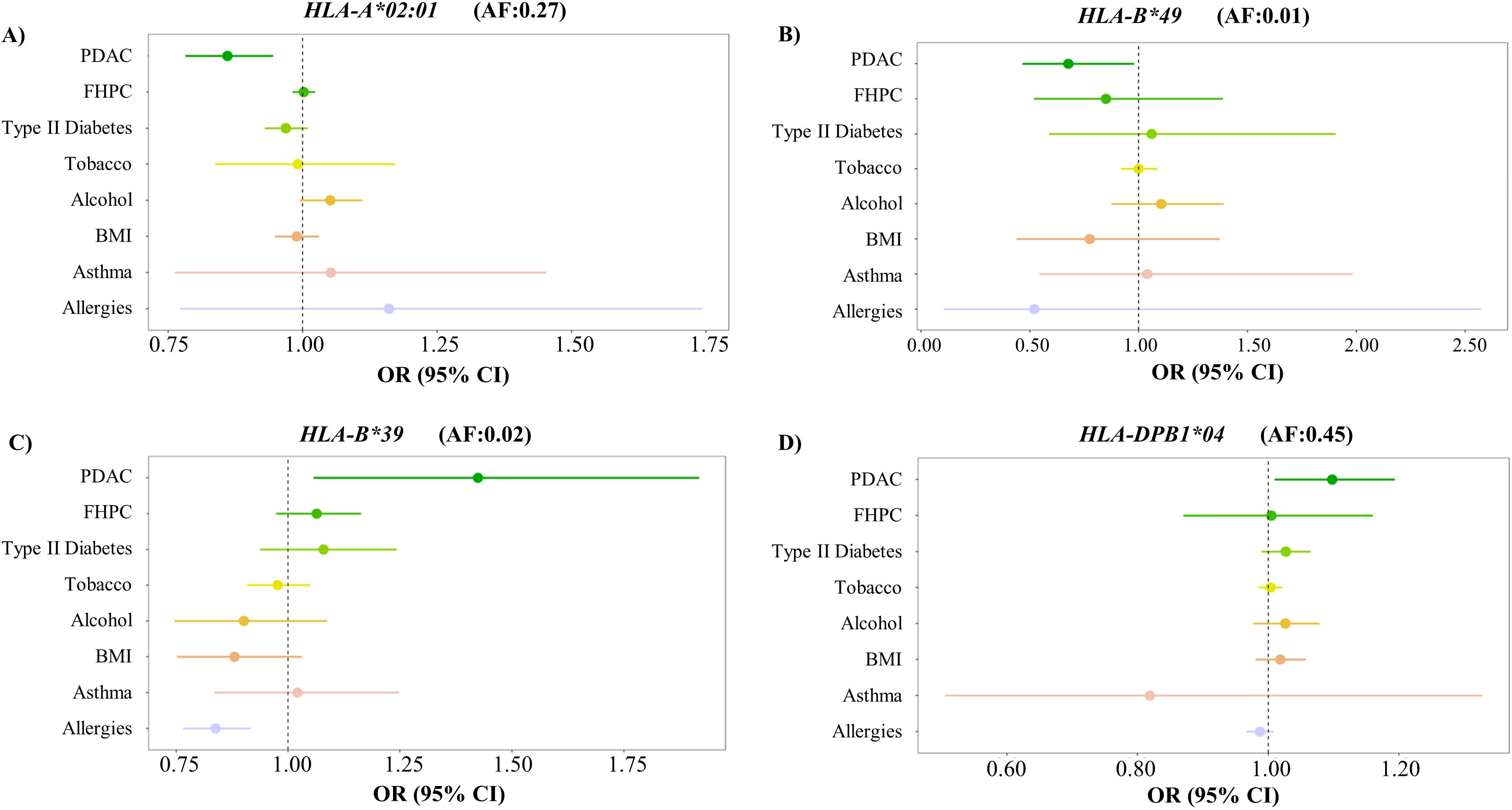
Odds ratio (OR) and 95% confidence interval (CI) of the association analyses between PDAC susceptibility alleles, as well as the allele frequency (AF): *HLA-A*02:01* (Figure 2A), *HLA-B*49* (Figure 2B), *HLA-B*39* (Figure 2C), and *HLA-DPB1*04* (Figure 2D) obtained in the meta-analysis and known risk/protective factors for PDAC.

Furthermore, the PanGenEU study identified one allele significantly associated with PDAC: the *HLA* class I *A*26:01* (OR=1.75, corrected p-value=4.17×10^−2^). Individuals carrying this allele, present in 5% of the PanGenEU study population, showed a higher risk of developing PDAC (OR=1.74, **Supplementary Table S4**). Additionally, when both controls and cases were combined to increase statistical power, we found that this allele was significantly and inversely associated with nasal allergies (OR=0.56, p-value=2.87×10^−2^) and, although non-significantly, with asthma (OR=0.68, p-value=2.2×10^−1^, **Supplementary Fig. S4A**). However, we did not replicate the association between *HLA-A*\*26:01 and PDAC risk in the UKB study (**Supplementary Table S5**) nor the meta-analysis, although the direction of the association was the same (OR=1.07, p-value=9.88×10^−1^).

### Cumulative PDAC risk score for the most promising alleles

We estimated cumulative PDAC risk by combining known risk and protective factors and adding PDAC-associated *HLA* alleles. In a meta-analysis with the UK Biobank, individuals with the abovementioned non-genetic high-risk conditions had 2.8 times higher PDAC risk. By including either *HLA-B*\*39 or *HLA-DPB1*\*04 alleles, the ORs increased to 5.28 and 3.34, respectively (**Table 1**), though not statistically significant (p-value=1.17×10^−1^, p-value=1.33×10^−1^, respectively). Conversely, asthmatic or allergic individuals who did not smoke, had no diabetes, nor had a family history of PDAC, experienced a 62% reduction in the odds (OR=0.38) of developing PDAC. Subjects who had one/two alleles of *HLA-A*\*02:01 further decreased their ORs to 0.32 and 0.27, respectively (p-value=1.18×10^−2^). By including one/two copies of the *HLA-B*49* allele in the model, the OR significantly decreased to 0.19 and 0.1 (p-value=2.4×10^−2^, **Table 1**). Additionally, in the PanGenEU study, diabetics, smokers, and individuals with a family history of cancer but without asthma or allergies had 4.33 times higher risk of PDAC than the population in the lowest risk category. When those individuals at high-risk harbor one or two copies of the *HLA-A*\*26:01 allele, the ORs increased to 7.73 and 13.81, respectively (p-value=2.28×10^−2^, see Supplementary Fig. S4B).

**Table 1.**
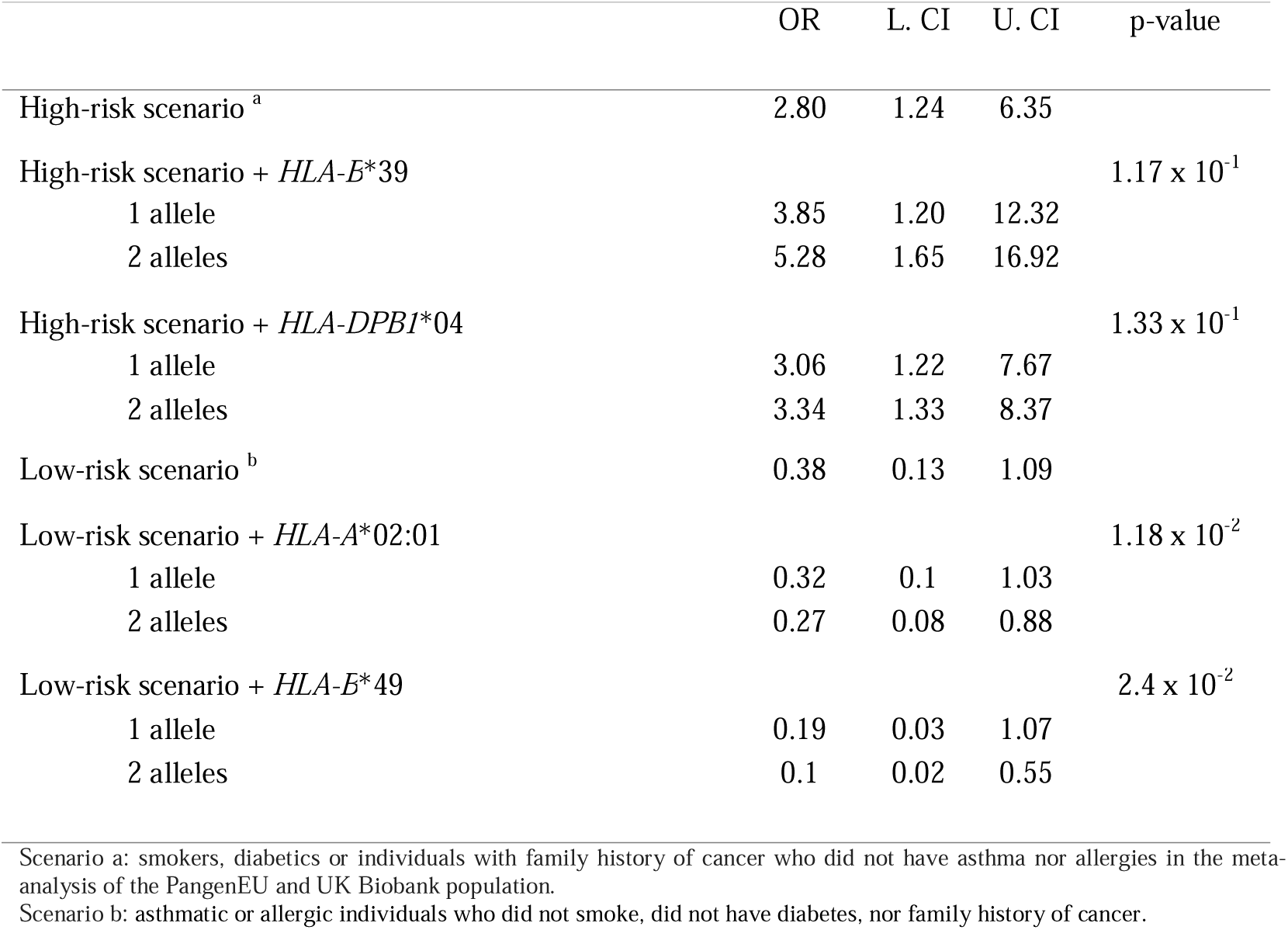
Cumulative PDAC risk estimates (OR: odds ratio and CI: confidence interval) in the extreme scenarios according to the presence/absence of known risk factors (Type II diabetes, smoking, and family history of pancreatic cancer, FHPC) or protective factors (asthma and nasal allergies) in addition to the significant *HLA* alleles identified in the meta-analysis.

### Association analyses between HLA haplotypes and PDAC risk

Given the critical role of *HLA* haplotypes in immune responses, we assessed their association with PDAC risk. We identified 18 haplotypes significantly associated with PDAC risk, nine of which remained significant after multiple testing correction (**Supplementary Table S6**). Two of the top five protective haplotypes carried the *HLA-A*02* allele, present in 20% of the European population and associated with decreased PDAC risk in our analyses. Haplotypes harboring the *HLA-A*02* allele were all protective for PDAC, some reaching an OR∼0.4). Among the haplotypes with the lowest effect size *HLA-B*35* and *HLA-A*02* alleles were common: *HLA-B*35*/*HLA-A*0*2 (OR=0.41, 95%CI=0.18-0.94), *HLA-DRB1*13*/*HLA-B*35* (OR=0.43, 95%CI=0.38-0.5), and *HLA-DRB1*01*/*HLA-A*02* (OR=0.49, 95%CI=0.27-0.89). On the contrary, the highest risk haplotypes harbored the *HLA-DRB1*04* allele: *HLA-DRB1*04*/*HLA-B*44*/*HLA-C*07* (OR=1.19, 95%CI=1.12-1.26) and *HLA-DRB*04*/*HLA-B*14* (OR=1.28, 95%CI=1.19-1.38) (**Supplementary Table S6**).

The *HLA-A*26*, *HLA-B*39*, and *HLA-B*49* alleles were not part of the haplotypes significantly associated with PDAC risk in the meta-analysis, probably because of their low AF: *HLA-A*26*=0.02, *HLA-B*39*=0.02, and *HLA-B*49*=0.01.

### HLA alleles are associated with overall survival of PDAC patients

We further assessed the role of *HLA* alleles in PDAC prognosis through a meta-analysis of the results from the survival analyses in PanGenEU, UK Biobank, and TCGA studies. 156 *HLA* class I and II alleles overlapped between the three studies (**Supplementary Fig. S5**). The results obtained under a dominant MoI revealed that patients who carried the *HLA-A*02:01* allele presented better survival than non-carriers (HR=0.87, p-value=3.3×10^−2^, **Fig. 3**), indicating that a single copy of this allele is enough to improve survival. On the other hand, homozygous individuals for *HLA-DPB1*04* displayed a borderline significantly worse survival under a codominance model (HR=1.19, p-value=6.3×10^−2^, **Supplementary Fig. S6**). Additionally, carriers of *HLA-A*30:01* (HR=1.75, p-value=1.2×10^−2^), and *HLA-DPB1*09:01* (HR=3.36, p-value=1.5×10^−3^) were also associated with worse overall survival compared to non-carriers (**Supplementary Fig. S7**).

**Figure 3.**
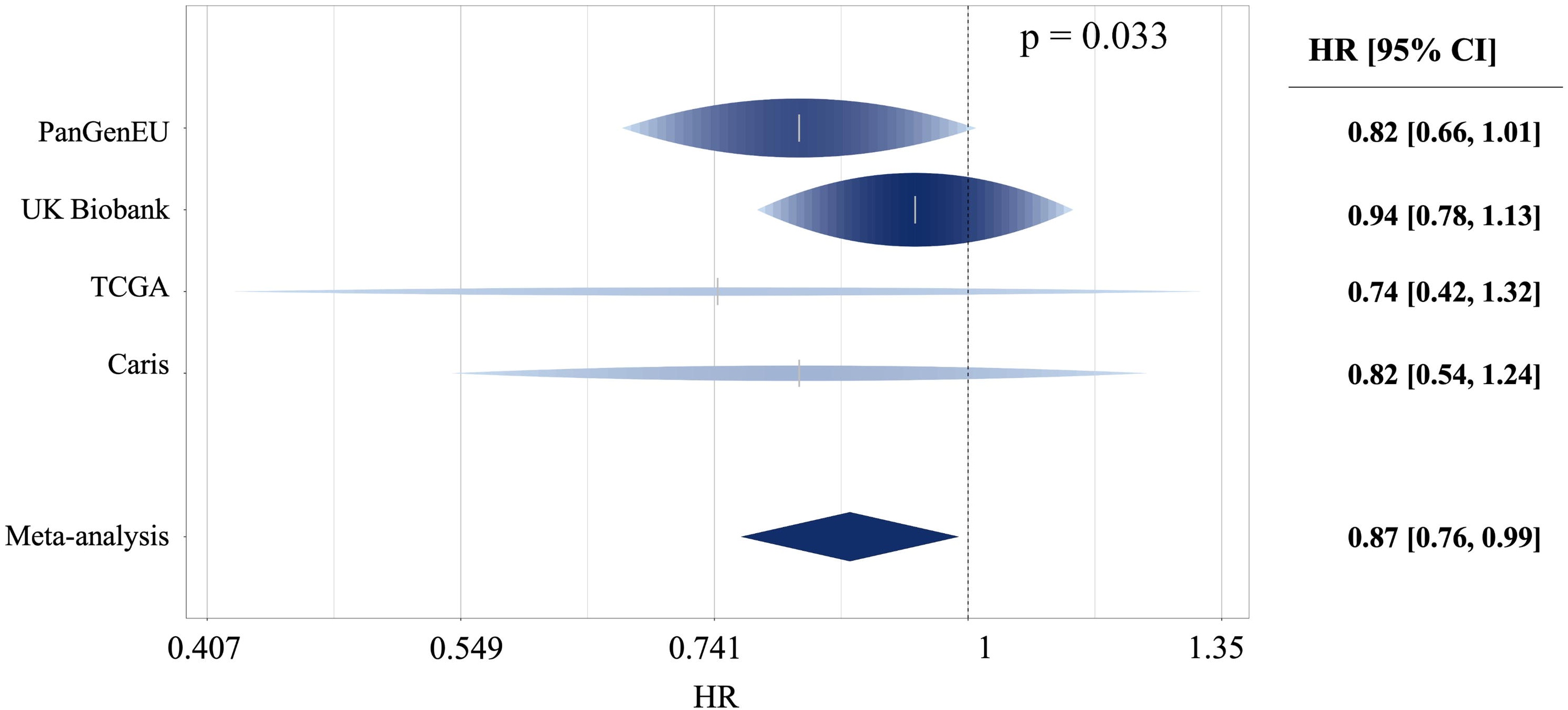
Hazard ratio (HR) and 95% confidence interval (CI) of the Cox proportional-hazards regression models under a dominance MoI obtained in the meta-analysis for the allele *HLA-A*02:01* when considering PanGenEU, UK Biobank, and TCGA (Figure 3A), and when including data from the Caris clinical trial in the meta-analysis (Figure 3B).

### HLA-A*02:01 interacts with KRAS^G12V^ mutations for PDAC survival

More than 90% of PDAC harbor *KRAS* mutations. We found that the PDAC protective allele *HLA-A*\*02:01 was associated with tumors harboring *KRAS*^G12V^ vs. other mutations in PDAC, NSCLC, and colorectal cancers (OR=1.55, p-value=4.65×10^−2^), and with *KRAS*^G12D^ mutations in gastric tumors (OR=7.27, p-value=6.42×10^−3^) (**Supplementary Table S7**).

Stratified analysis revealed that *HLA-A*02:01* carriers with *KRAS*^G12V^ tumors had significantly better survival (HR=0.19, p-value=1.3×10^−2^, **Fig. 4A**) than *HLA-A*\*02:01 carriers with other *KRAS* mutations; importantly, survival did not differ for patients with *KRAS*^G12V^ mutated tumors who did not carry the *HLA-A*02:01* allele (HR=0.87, p-value=8.1×10^−1^, **Fig. 4B**). Conversely, carriers of *HLA-A*02:01* and *KRAS*^G12D^ had significantly worse survival (HR=5.51, p-value=1.5×10^−3^, **Fig. 4C**) than non-carriers (HR=1.13, p-value=6.2×10^−1^, **Fig. 4D**). Further stratified analyses focusing on individuals carrying either the *KRAS*^G12V^ or *KRAS*^G12D^ mutations, along with the presence or absence of the *HLA-A*02:01* allele, showed that the longer survival of *KRAS*^G12V^ carriers appear to be driven by the interaction with the *HLA-A*02:01* allele (HR=0.13, p-value=4.4×10^−2^, **Fig. 5A**). However, for those with *KRAS*^G12D^ mutated tumors, the decreased survival was mainly due to the mutation itself, regardless of the *HLA* allele (HR=1.21, p-value=2.2×10^−1^, **Fig. 5B**). Interestingly, we discovered that *KRAS*^G12D^ mutated tumors, displayed a significant higher expression level of IFN-γ in comparison to tumors carrying any of the other *KRAS* mutations (**Supplementary Fig. S8**).

**Figure 4.**
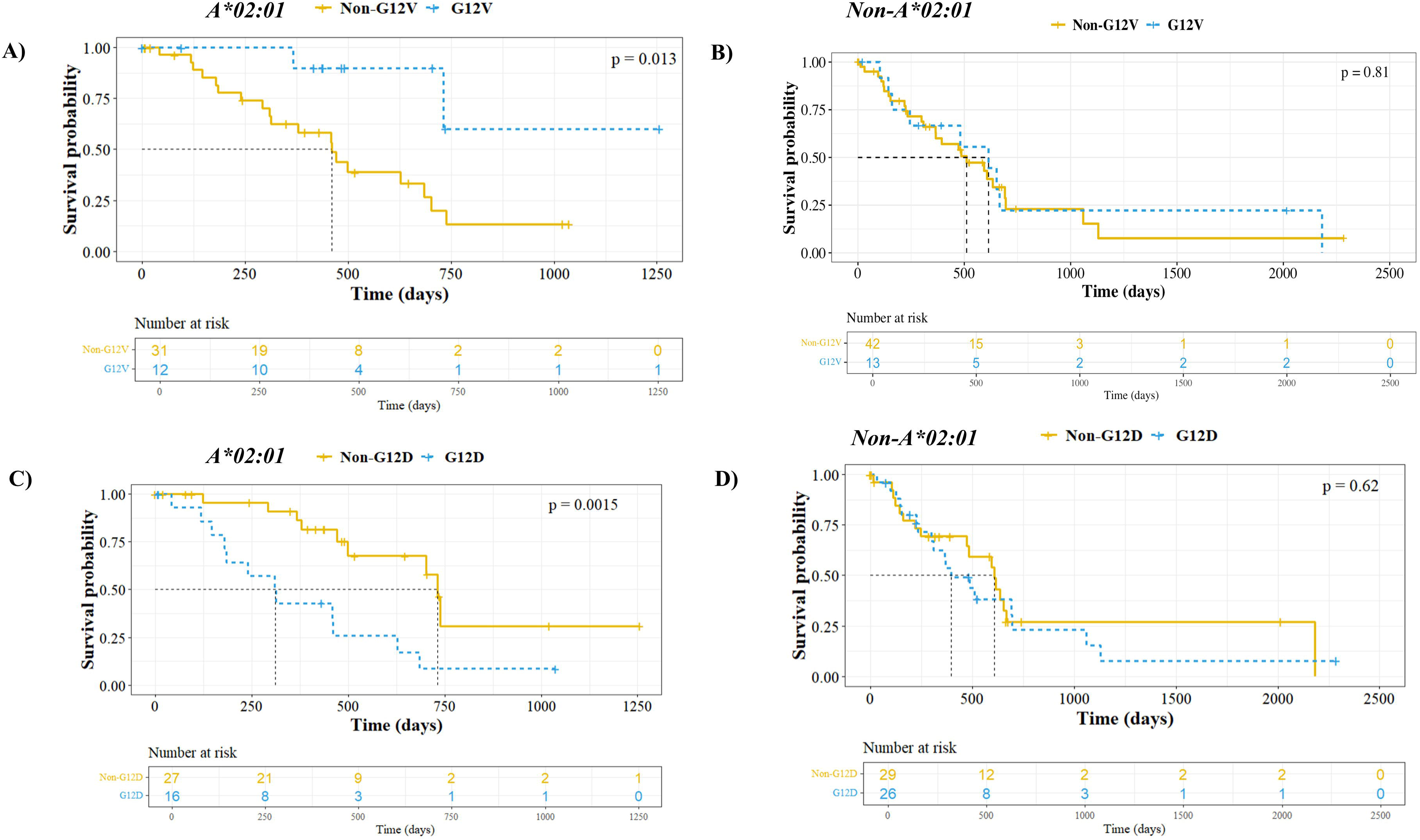
Kaplan-Meier curves of patients according to their KRAS mutational status in *HLA-A*02:01* carriers with PDAC tumors harboring *KRAS*^G12V^ patients whose tumors have other KRAS mutations who are carriers of *HLA-A*02:01* (Figures 4A and Figure 4C) and non-carriers of that allele (Figures 4B and Figure 4D).

**Figure 5.**
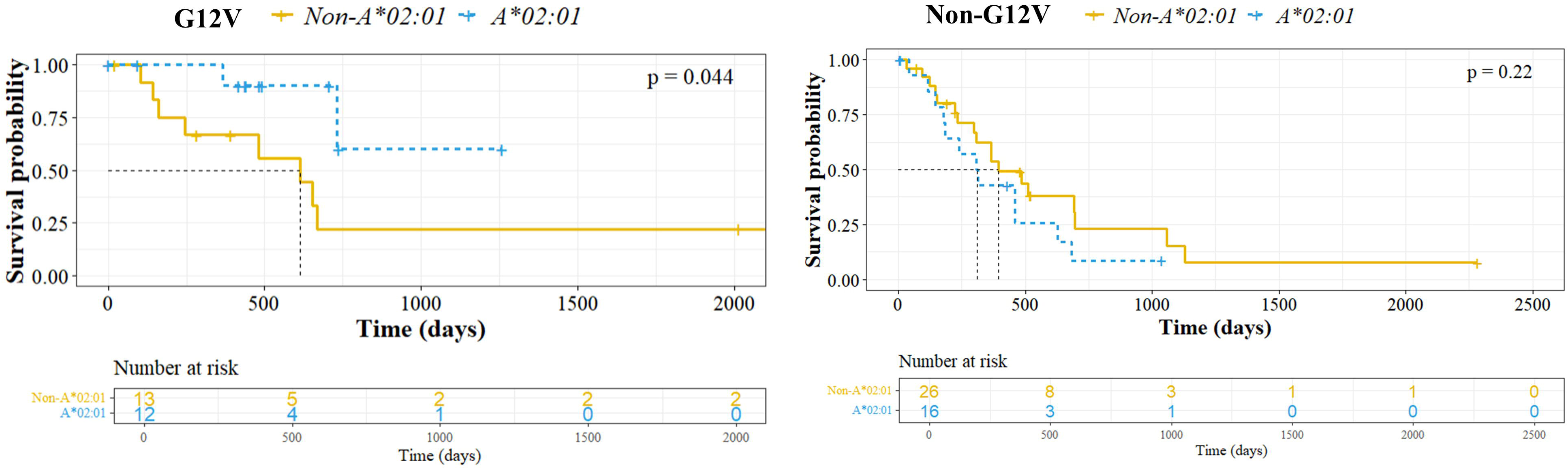
Kaplan-Meier curves of patients according to the presence or absence of the *HLA-A*02:01* allele in *KRAS*^G12V^ (Figures 5A) and *KRAS*^G12V^ (Figures 5B) mutated tumors.

The interaction model in the TCGA study showed that carriers of the *HLA-A*02:01* allele who had a *KRAS*^G12V^ mutated tumor had a significantly lower risk of death (HR=0.14, p-value=2.4×10^−2^) compared to patients with any of the other *KRAS* mutations, adjusted for gender, age at diagnosis, stage, and Moffit tumor subtype. We then performed a meta-analysis of the interaction between the *A*02:01* allele and *KRAS*^G12V^ mutations, including the PAN-NGS and Caris populations (**Supplementary Fig. S9**). This interaction held significant in early-stage tumors (HR=0.25, p-value=1.7×10^−2^, **Fig. 6A**) but not in advanced-stage (HR=0.93, p-value=8.7×10^−1^, **Fig. 6B**).

**Figure 6.**
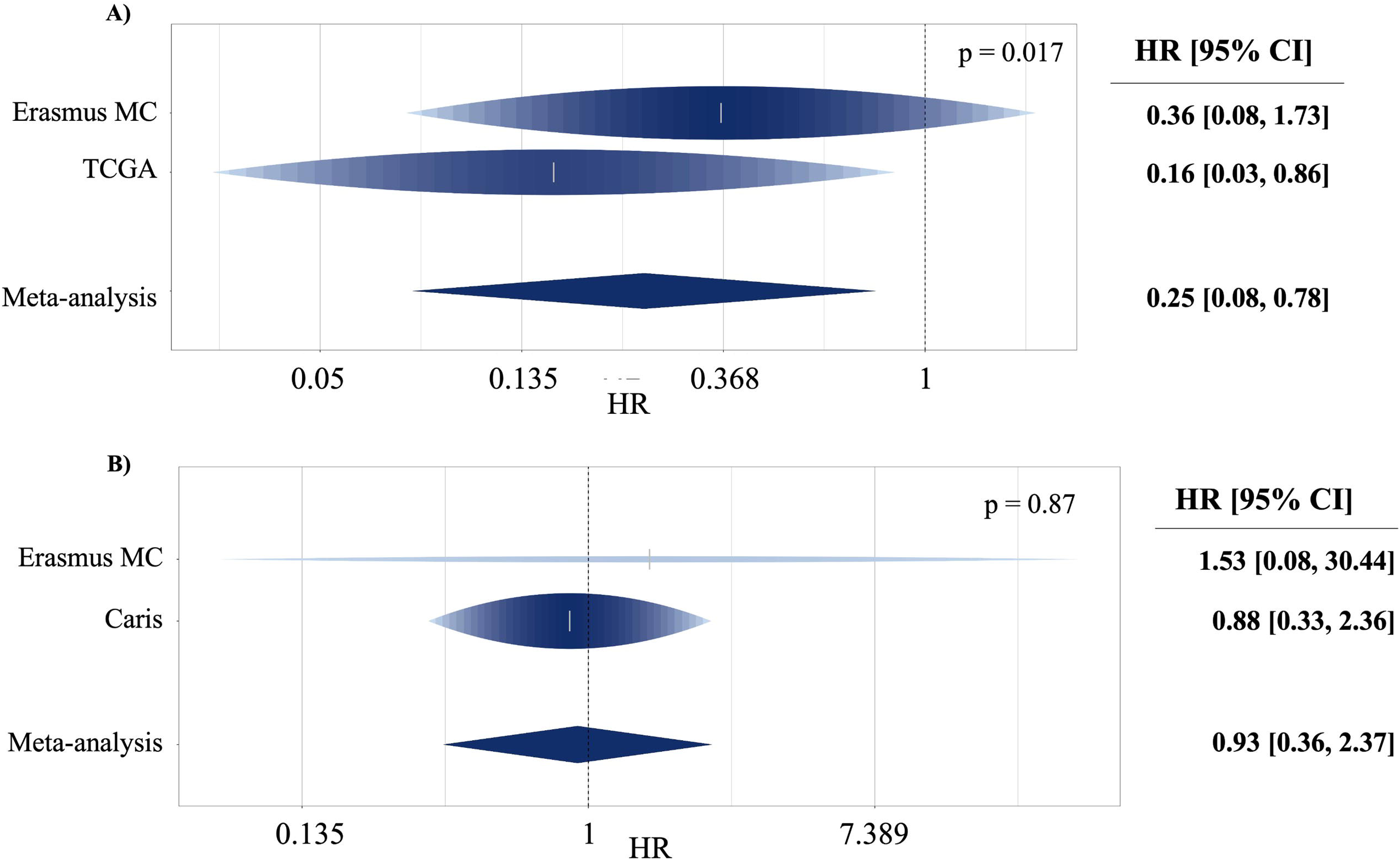
Hazard ratio (HR) and 95% confidence interval (CI) of the Cox proportional-hazards regression models under a dominance MoI obtained in the meta-analysis for the allele *HLA-A*02:01* for (A) stages I&II PDAC tumors from Erasmus and TCGA study populations and (B) stages III&IV tumors from Erasmus MC and Caris study populations.

## DISCUSSION

The present study represents the most exhaustive analysis of the role of the *MHC* region at the *HLA* allele and haplotype levels associated with PDAC risk and prognosis. We show that different *MHC* genetic profiles recognize distinct PDAC risk populations. We identified five novel susceptibility alleles in the *HLA* class I and II regions associated with PDAC risk: *HLA-A*\*26:01, *HLA-A*\*02:01, *HLA-B*\*49, *HLA-B*\*39, and *HLA-DPB1*\*04. Among these, *HLA-A*02:01* remained significant after correcting for multiple testing, showing a notably reduced risk of PDAC (metaOR=0.86, corrected p-value=2.25×10^−2^. This allele is prevalent in 20% of the European population, and was present in all the protective haplotypes. Interestingly, HLA-A*02:01 has been found to be associated also with a decreased risk of melanoma (29). Importantly, we found that germline *HLA-A*02:01* carriers with PDAC from the PanGenEU, UKB, TCGA, and Caris had a survival benefit (metaHR=0.87, p-value=3.3×10^−2^; **Fig. 3B**). These findings go along with the observations that *HLA-A*02:01* carriers have better survival in NSCLC (30), and might have improved outcomes to immunotherapy (31), pointing to *HLA-A*02:01* as a good prognostic marker for PDAC. Notably, the *HLA-A*02:01* allele is among the eligibility criteria in different cancer immunotherapy clinical trials for esophageal carcinoma (32), acute myeloid leukemia (33), NSCLC (34), and glioblastoma (currently in phase III) (35).

Since *HLA-A*02:01* has shown a high predicted binding affinity for the *KRAS* codon 12 mutation in silico-in vitro studies (36), we also assessed the association of *HLA-A*02:01* with the most common hotspot *KRAS* mutations (at codons 12, 13, and 61) in *KRAS*-mutated cancers (PDAC, NSCLC, stomach, and colorectal) using TCGA data. After adjusting for cancer type, we discovered that *HLA-A*02:01* is preferentially associated with tumors harboring *KRAS*^G12V^ mutations (OR=1.55, p-value=4.65×10^−2^). Remarkably, we observed that patients with *KRAS*^G12V^ mutated tumors who carried the *HLA-A*02:01*, comprising approximately 5% of the PDAC patients, experienced significantly better survival compared to those without the allele. This novel interaction was confirmed in an independent series of resectable (or early-stage) PDAC cases (PAN-NGS). Supporting this host-tumor interaction, the NEPdb (https://nep.whu.edu.cn/) database, which includes data on over 17K validated human immunogenic neoantigens, shows that only *KRAS*^G12V^ and *KRAS*^G12D^ mutations have been validated to bind the *HLA-A*02:01* protein and elicit an immune response. Conversely, regardless of the HLA allele, patients with *KRAS*^G12D^-mutant tumors had significantly worse survival than those without this mutation. However, the PAN-NGS and Caris independent datasets could not validate this finding.

Therefore, this study is a pioneer in showing that the presence of *HLA-A*02:01* allele not only is associated with protection against PDAC risk but it is also associated with better PDAC survival, mainly in patients with early-stage *KRAS*^G12V^ mutant tumours. Our findings could lead to an *HLA-A*02:01*-*KRAS*^G12V^ classification system for nearly 2% of PDAC patients with better survival. Also, we anticipate that these patients may benefit from immunotherapy based on the results observed in xenograft models, with the *HLA-A*02:01* showing a better response to immunotherapy due to its ability to bind *KRAS*^G12V^ neoantigens (31). In contrast, approximately 40% of PDAC patients with *KRAS*^G12D^ mutated tumors demonstrated the poorest survival outcomes in TCGA, independent of their *HLA* allele. This significantly narrows the therapeutic options for these patients, potentially restricting them to systemic chemotherapy alone. Given that *KRAS* mutations are neoantigens capable of binding the *HLA-A*02:01* protein and initiating an immune response, we hypothesize that *HLA-A*02:01*-*KRAS*^G12V^ individuals might have an enhanced immune response characterized by increased infiltration of anti-tumor immune cells, particularly CD8^+^ T cells, compared to those with the allele and another *KRAS* mutation. Conversely, patients with *KRAS*^G12D^ mutated tumors might either produce less immunogenic neoantigens or exhibit a higher infiltration of cells able to promote tumor tolerance. This results in a reduced ability of cytotoxic T cells to recognize and target the tumor cells. Interestingly, we discovered that *KRAS*^G12D^ PDAC tumors exhibited higher expression levels of IFN-γ in comparison to other *KRAS* mutated tumors. The differences in survival that we found in *KRAS*^G12D^ PDAC tumors might be due to elevated IFN-γ levels upregulate PD-L1 expression, which binds to PD-1 receptors on T cells, inhibiting their activity and the recruitment of regulatory T cells (Tregs) and myeloid-derived suppressor cells (MDSCs) to the tumor microenvironment (37). In addition, high IFN PDAC tumors have been associated with worse overall survival, due to a higher activation of stroma contributing to tumor growth (38). Altogether might explain the differences in survival that we found in *KRAS*^G12D^ PDAC tumors. Consistently, a recent study published by Till et al., 2024, demonstrated that the clearance of ct*KRAS*^G12D^ is associated with improved overall outcomes in metastatic PDAC. This might be due to the immune system no longer recognizing the G12D neoantigen, leading to reduced IFN-γ levels (39).

Beyond A*02:01, we identified three additional alleles associated with PDAC risk in our meta-analysis. Among them, *HLA-DPB1*04* was associated with an increased risk of PDAC, with haplotypes containing this allele also showing an increased risk. Limited information is available on the role of this allele in cancer (40). Previous studies have shown that the risk allele *HLA-B*39* is associated with Grave’s disease (41) while the protective allele *HLA-B*49* has been inversely associated with COVID-19 (42). Nevertheless, our study is the first to report an association between these alleles and cancer risk. Finally, *HLA-A*26:01* was associated with an increased risk of PDAC and inversely associated with nasal allergies and asthma in the PanGenEU population. These results point to a possible explanation of previous findings showing an inverse association between atopic comorbidities and PDAC risk (10). While the meta-analysis combining PanGenEU and UKB data threw a risk estimate (OR=1.07), it lacked statistical significance. This could be accounted for by the low AF of *HLA-A*26:01* in UKB population (0.02) vs the AF in PanGenEU (0.06), the latter AF being similar in most European countries with UK differing in the AF by >50%. *HLA-A*26:01* has been reported to be associated with an increased risk of non-Hodgkin lymphoma (OR=8.5, p-value=2.6×10^−2^) (43). Importantly, including these alleles in a non-genetic-based cumulative risk score significantly altered the risk of developing PDAC, either increasing or decreasing it substantially.

Our study has some limitations. First, the analyses employed self-reported epidemiological information from subjects included in the studies that could lead to misclassification of the risk factors and comorbidities, such as asthma and rhinitis. However, our meta-analysis using the estimates of both PanGenEU and UKB populations allowed us to minimize this potential bias. In addition, we restricted the analyses to the Caucasian population; therefore, the impact of *HLA* alleles on PDAC risk and prognosis in other ethnic groups should be further assessed. Additionally, the *MHC* region is one of the most polymorphic areas of the human genome, making it challenging for association analyses to detect rare alleles that may play a crucial role in PDAC risk.

Despite the above-mentioned limitations, our work has several strengths. We performed an in-depth analysis of the relationship between the *HLA* region and PDAC risk by employing state-of-the-art methodology using important resources from two large and independent study populations with complementary designs: case-control and cohort. We corrected for population stratification by adjusting for the PCAs. Additionally, the association with PDAC prognosis was first conducted with PanGenEU study and further validated with data from three additional independent datasets: UKB, TCGA, PAN-NGS, and Caris.

Our findings provide a strong basis for *HLA-A*02:01*, a single biomarker to be considered in improving PDAC prevention and risk stratification, allowing for identifying patients who may require more aggressive monitoring or treatment due to their lower genetic risk of mortality. These latter results warrant improving clinical trials based on *HLA* allele and *KRAS* mutation profiling. Future studies should also include PDAC subtypes in the association analysis and extend the scope to other *KRAS*-related cancer types. Since tumors can lose the expression of the *HLA* alleles, we suggest assessing whether this loss is happening in specific alleles with a protective role. Finnaly, given the crucial role of *HLA* alleles in immune responses, we suggest evaluating the immunotherapy response in PDAC patients according to their *HLA* allelic profile.

## MATERIAL AND METHODS

### Ethics Statement

IRB ethical approval and written informed consent were obtained by all participating centers contributing to PanGenEU, and study participants, respectively. The study was conducted in accordance with the Helsinki Declaration.

### Study populations

In this study, we employed the resources of the PanGenEU case-control study, the UKB cohort, and datasets from The Cancer Genome Atlas (TCGA), PAN-NGS trial from Erasmus Medical Center (EMC), Rotterdam, and Caris trial from Vall d’Hebron University Hospital (VHIO), Barcelona.

*PanGenEU study* details have been reported elsewhere (9–11,44). It is a multicentric case-control study on PDAC conducted in 28 centers across Spain, Italy, Sweden, Germany, the UK, and Ireland from 2009-2014 and 2016-2018. Newly diagnosed PDAC patients over 18 years of age were recruited. Eligible controls were hospital inpatients with primary diagnoses not associated with any of the well-known PDAC risk factors. Controls for Ireland and Sweden were population-based. Epidemiological information was collected through direct in-person interviews by trained health monitors using structured questionnaires, including information about lifestyle, environmental exposures, and medical history (9). The individuals were genotyped with the Infinium OncoArray-500K (45) and missing genotypes were imputed using IMPUTE2 v2 (46) and 1000Genomes (Phase 3, v1) as reference (47). Quality control filters included the missing call rate, unexpected heterozygosity, discordance between reported and genotyped gender, unexpected relatedness, and estimated European ancestry < 80%. Once those samples were filtered out, genotyped data from 1,317 cases and 700 controls were employed for *HLA* allele imputation and association analyses.

*UKB* is a prospective cohort study of approximately 500,000 individuals from the United Kingdom (48). The UKB Axiom array was used for genotyping 450,000 subjects, and the remaining 50,000 individuals were genotyped with the UK BiLEVE Axiom array. Based on their primary International Classification of Diseases (ICD9 and ICD10) codes, we selected 935 PDAC patients (**Supplementary Table S8**). We discarded the self-reported cases (N=45) to ensure PDAC diagnosis reliability and those individuals with no Caucasian genetic ethnicity (N=129), resulting in a final set of 761 PDAC cases. After selecting Caucasian individuals without a history of cancer and with the same ICD9 and ICD10 codes as PanGenEU controls (10), the final dataset of controls included 95,050 subjects.

*TCGA*, established in 2006 by the National Cancer Institute and the National Human Genome Research Institute, is one of the largest databases of cancer genomics data (49). TCGA has genetic information for 33 cancer types, genotyped with the Affymetrix 6.0 SNP array. We collected information on outcome, *HLA* alleles typed with the Optitype tool (50), and *KRAS* mutations from PDAC, colorectal, stomach adenocarcinoma, and Non-Small Cell Lung Cancer (NSCLC) patients. We kept for the analysis Caucasian individuals and those patients with complete data, 101 PDAC, 185 colorectal, 196 stomach adenocarcinoma, and 620 NSCLC. Additionally, we employed data from Thorsson et al, 2018 to assess the immune landscape of the 101 TCGA PDAC tumors.

The PAN-NGS trial (CCMO ID NL75415.078.20) includes PDAC patients from twelve centers throughout the Netherlands who were prospectively recruited from April 2021 to June 2023. Local review committees approved the trial. Eligible patients had cytologically or histologically confirmed PDAC, were aged 18-60 years at diagnosis, had an Eastern Cooperative Oncology Group performance score of 0-2, and had an estimated life expectancy of at least twelve weeks. Patients were excluded when they did not want to know if alterations might be associated with genetic predisposition of cancer or in case of locally advanced or local recurrence of PDAC without histological tissue readily available. Clinical data were collected at participating centers and supplemented as needed. Clinical information was obtained at the site of the participating centers, and missing information was requested if were also treated in other hospitals. A final dataset of 108 PDAC patients was employed for the subsequent analysis.

*Caris clinical trial* is a retrospective single-center dataset involving patients with PDAC treated at Vall d’Hebron University Hospital between 2021 and 2023. Eligible participants were over 18 years old, had histologically confirmed PDAC, and underwent molecular analysis with the Caris Life Sciences sequencing panel. Exclusion criteria were non-adenocarcinoma pancreatic tumors, poor-quality samples unsuitable for sequencing, inadequate follow-up, or lack of written informed consent. Clinical data were collected at diagnosis, and a multidisciplinary team assessed the clinical stage. The Ethics Committee of the Vall d’Hebron University Hospital (PR(AG)651/2023) approved the study. Complete data was obtained for 145 PDAC patients and included in the analysis.

### HLA (or MHC) alleles assessment

*HLA* class I and II alleles were first imputed using genotyped data from the PanGenEU study. We took advantage of the imputed *HLA* alleles from UKB, TCGA, Erasmus MC, and Caris subjects.

#### PanGenEU study

The imputation was done by *SNP2HLA* software (version v 1.0.3) (51) considering 5,790 SNPs within the *MHC* region with an info parameter ≥0.9. This info parameter ranges from 0 to 1 and assesses the quality/reliability for a particular variant imputed through IMPUTE2. We focused on SNPs overlapping with those from the Type 1 Diabetes Genetic Consortium (T1DGC) V1.0.3, the largest European reference panel (52). This dataset includes 5,225 individuals with genotyping data for 7,238 SNPs, 424 *HLA* alleles (126 1-field and 298 2-field alleles), and 1,276 HLA amino acids. It allowed an accurate imputation of classical *HLA* alleles and amino acids of class I (*HLA-A*, -*B*, and -*C*) and class II (*HLA-DPB1*, - *DPA1*, -*DQB1*, -*DQA1*, and -*DRB1*) genes. We excluded *HLA* alleles that were not in Hardy-Weinberg equilibrium (HWE) in controls (p-value<10^−6^), calculated using the *HardyWeinberg* R package V1.6.3 (53), those with a bad imputation quality (info<0.3), as well as those with an allele frequency (AF) <5%, ending up with 40 alleles of 1-field, and 39 alleles of 2-fields.

*UKB* information on 362 *HLA* alleles across eleven loci (*HLA-A*, -*B*, -*C*, -*DPB1*, -*DPA1*, -*DQB1*, -*DQA1*, -*DRB1*, -*DRB2*, -*DRB3*, -*DRB4*, and -*DRB5*) was available and obtained through the HLA*IMP:02 program (54).

#### TCGA

*HLA* class I alleles from the colorectal, stomach, and NSCLC tumors were inferred from normal WES using polysolver (55). Then, xHLA (56) was employed to infer the *HLA* I and *HLA* II alleles (*HLA-A*, -*B*, -*C*, -*DPB1*, -*DQB1*, and - *DRB1*) from normal WES of the PDAC patients.

#### Erasmus MC

*HLA* genotype results were determined based on information from mRNA isolated from a formalin-fixed paraffin-embedded tumor sample using the Agilent SureSelectXT Low Input Library prep chemistry, optimized for FFPE tissue, in conjunction with the SureSelect Human All Exon V7 bait panel (48.2 Mb) and the Illumina NovaSeq.

#### Caris clinical trial

HLA genotypes were obtained from Caris Life Sciences reports for all patients. Genomic DNA was isolated from microdissected, formalin-fixed paraffin-embedded tumor samples and analyzed using the Illumina NovaSeq 6000 sequencers. A hybrid pull-down panel of baits designed to enrich for more than 700 clinically relevant genes at high coverage and high read-depth was used, along with another panel designed to enrich for additional >20,000 genes at a lower depth. To assist with gene amplification/deletion measurements and other analyses, a 500Mb SNP backbone panel (Agilent Technologies) was included. Analytical validation for *HLA* genotyping showed >99% concordance with a validated comparator method.

### HLA haplotype assessment

We inferred the haplotypes independently in each population using the *haplo.stats* R package (57) after phasing with BEAGLE (58) and considering the *HLA* gene combinations prevalent in the Net Allele Database (28). A final set of 3,129 haplotypes were inferred in the PanGenEU individuals using 262 1-field *HLA* alleles across all eight loci with an info parameter reflecting imputation quality >0.3. In the UKB study, a final set of 148 and 57 1-field *HLA* class I and *HLA* class II haplotypes, respectively, were identified using 141 1-field *HLA* alleles. No *HLA* haplotype assessment was done for the TCGA population.

### Statistical analyses

We assessed the association between the *HLA* region and PDAC risk for the individual *HLA* alleles and haplotypes within each study population. Subsequently, we combined the summary statistics using a meta-analysis approach. Then, we studied the association of the *HLA* alleles with well-established PDAC risk factors. Additionally, we investigated the heritability explained by *HLA* class I and class II alleles (**Fig. 1**). We did not consider further the analysis at the amino-acid level because significant results at the individual level were not found (**Supplementary Table S9**). Finally, prognostic analyses integrated *HLA* alleles and *KRAS* mutational profiles.

#### HLA allele-based association analysis with PDAC risk

Association analyses were done using the *PLINK* V1.9 software (59). In the PanGenEU setting, we tested the association between the *HLA* alleles and PDAC risk at the individual level by estimating the Odds Ratios (OR) and 95% Confidence Intervals (CI) through logistic regression models including age, gender, the European region, and the first five principal components (PCAs) to correct for population stratification. We considered the three Modes of Inheritance (MoI) for 424 PanGenEU *HLA* class I and II alleles with AF >0.05. In UKB, we included the I- and II-fields *HLA* alleles with an AF >0.1% (60,61). We performed logistic regression models adjusting for the same covariates as in the PanGenEU models, plus the type of genotyping array and center of inclusion.

A posterior meta-analysis using a random-effects model was performed with the *rma.uni* function of the *metafor* R package (62), combining the risk estimates of *HLA* alleles for PDAC with a nominal p-value<0.1 from the association analysis in PanGenEU and UKB studies, followed by multiple testing correction.

#### HLA allele-based association analysis with established PDAC risk factors

We used logistic regression models to assess the association between *HLA* alleles significantly associated with PDAC risk in the previous meta-analyses with established PDAC risk factors such as Family History of Pancreatic Cancer (FHPC), T2DM, obesity, and tobacco and alcohol consumption, as well as with PDAC protective factors (asthma and nasal allergies). We restricted these analyses to the controls from each study. Then, we performed a meta-analysis by applying a random-effects model. A cumulative PDAC risk score for the most promising alleles was computed by using the estimates obtained in the meta-analysis including PanGenEU and UKB.

#### Haplotype association analysis

We used the *haplo.stats* R package (version 1.7.9) to perform the statistical association analysis of the PanGenEU and UKB *HLA* haplotypes with PDAC risk. We employed a logistic regression model adjusted for the same covariates as in the previous analyses. We considered the three MoIs in each study population. Multiple testing correction was applied for each gene combination separately after discarding the low prevalence *HLA* haplotypes (i.e., frequencies either <5% or <0.1% in PanGenEU and UKB, respectively).

### Estimation of the heritability explained by HLA class I and class II alleles

We applied a Bayesian multi-kernel-based regression (63) to estimate the genetic variance explained by *HLA* class I and *HLA* class II alleles using the resources of the PanGenEU study:

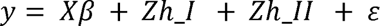

where the response variable *y* is a vector of case/control outcomes, ***X*** is an incidence or design matrix relating the adjusting covariates to each individual, p is the corresponding regression coefficient, ***Z*** is a design matrix allocating records ((***y***) to the vector of *HLA* class I/II associated risk scores (*h*_1_ and *h*_11_), and ε is a vector of random normal deviates with null mean and variance 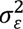 that includes model misspecification and environmental effects not considered in this model. The two genetic and residual variances were assumed to follow the distributions 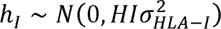, 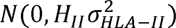 and 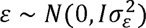, where *H*_1_ and *H*_11_ correspond to the *HLA* class I and *HLA* class II alleles-derived relationship matrices between individuals, 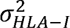 and 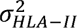 are the genetic variances explained by *HLA* class I and II alleles, **I** is an identity matrix, and 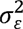 corresponds to the residual variance. The prior distribution for the genetic and residual variances were inverted chi-square distributions.

*H*_1_ and *H*_11_ similarity matrices were constructed using a Gaussian kernel with the *G.matrix* function of *snpReady* R package (64). Estimates of the unknown parameters were obtained from their posterior distributions using a Gibbs sampling implemented in the *BGLR* R package. We ran a McMC chain of 500,000 iterations, and the first 100,000 were discarded as burn-in.

#### HLA allele-based survival analysis with PDAC risk

We assessed patient survival based on the presence or absence of the *HLA* alleles, with a strong prior on *HLA-A*02:01*. To this end, we used the survival data of 425 PDAC cases from PanGenEU, 506 from UKB, 101 from TCGA (98 when we account for the stage of the tumor), and 145 from the Caris clinical trial. First, we assessed the overlap of the *HLA* class I and II alleles between the three study populations. We computed Hazard Ratios (HR) and 95%CI using Cox proportional-hazards regression models for each population separately under (co-)dominance MoIs. Adjustment covariates in PanGenEU models were sex, age at diagnosis, center, tumor stage, and the first PCAs; for UK Biobank, we included sex, age at diagnosis, genotyping array type, recruitment center and the first five PCAs; for the TCGA and Caris models, we considered sex, age at diagnosis, and tumor stage. Finally, we meta-analyzed the results from the four populations using the function *rma.uni* from the *metafor* R package.

#### Association analysis between HLA-A*02:01 and KRAS common mutations

Based on previous reports pointing to a joint effect of *HLA-A*02:01* and *KRAS* mutations (31,36), we further explored the interaction between these biomarkers using TCGA data. We first assessed the association between *HLA-A*02:01* and well-established *KRAS* missense mutations at codons 12 (G12A, G12C, G12D, G12F, G12R, G12S, G12V), 13 (G13C, G13D), and 61 (Q61H, Q61K, Q61L, Q61R) using logistic regression models in tumors with high proportion of *KRAS* mutations (PDAC, colorectal, stomach, and NSCLC) (**Supplementary Table S10**). As the reference category, we used the non-G12V mutations for PDAC and colorectal cancer and the non-G12C for NSCLC. We initially assessed the association within each specific cancer type individually. Subsequently, we pooled the data from all cancer types into a single model, adjusting for cancer type. Finally, we used a Cox proportional-hazards regression model to evaluate whether the interaction between *HLA-A*02:01* and the specific *KRAS*^G12V^ and *KRAS*^G12D^ mutations was associated with overall survival in PDAC. The same covariates mentioned in the previous paragraph and the Moffit subtype of the tumor were employed to adjust these models. We used a Wilcoxon test to evaluate whether the associations found in overall survival could be explained by infiltrated immune components. Differences were considered significant at p-value<0.05.

To validate the interaction between the HLA-A*02:01 and the *KRAS* mutations, we used data from two independent populations, the Erasmus MC study and the Caris clinical trial. We analyzed the interaction between the *HLA-A*02:01* allele and the *KRAS*^G12V^ mutation in each population, adjusting for variables such as sex, age at diagnosis, tumor stage, and treatment for Caris, and the same variables plus center of inclusion for Erasmus MC. We then conducted a meta-analysis using a random-effects model. Finally, we examined the effect of the interaction within each population by stratifying patients based on tumor stage (stages I-II and stages III-IV) and conducted separate meta-analyses for these subgroups.

## DECLARATIONS

### Availability of data and materials

The datasets generated and/or analyzed during the current study are not publicly available due to the sensitive nature of genetic data but are available from the corresponding author upon reasonable request.

## Authors’ contributions

Study conception: NM, ELM. Design of the work: AL, RR, ELM, NM. Data acquisition: RTL, AC, CvE, TM, MH, MI, XM, JML, CM, JP, MO, VMB, AT, AF, LMB, TCJ, EFM, TG, WG, LS, IF, GS, LLC, JB, EC, LI, JK, BK, JM, DO, AS, WY, JY, FXR, NM, and rest of the PanGenEU Investigators. Data analysis: AL, LA, SS, ELM. Interpretation of data: AL, RR, FXR, ELM, NM. Creation of new software used in the work: No one. Drafting the work or substantively revising it: AL, RR, FXR, ELM, NM. Approval of the submitted version (and any substantially modified version that involves the author’s contribution to the study): ALL AUTHORS. Agreement on both to be personally accountable for the author’s own contributions and to ensure that questions related to the accuracy or integrity of any part of the work, even ones in which the author was not personally involved, are appropriately investigated, resolved and the resolution documented in the literature: ALL AUTHORS.

## Data Availability

All data produced in the present study are available upon reasonable request to the authors

## Acknowledgements

The authors thank the patients, coordinators, field and administrative workers, and technicians of the European Study into Digestive Illnesses and Genetics (PanGenEU) and TCGA studies. This research has been conducted using the UK Biobank Resource under Application Number 47884. This research was performed using resources generated by Type 1 Diabetes Genetics Consortium, a collaborative clinical study sponsored by the National Institute of Diabetes and Digestive and Kidney Diseases (NIDDK), National Institute of Allergy and Infectious Diseases (NIAID), National Human Genome Research Institute (NHGRI), National Institute of Child Health and Human Development (NICHD), and Juvenile Diabetes Research Foundation International (JDRF) and supplied by NIDDK Central Repository (NIDDK-CR). This manuscript was not prepared under the auspices of the T1DGC study and does not necessarily reflect the opinions or views of the T1DGC study, study sponsors, NIDDK-CR, or NIH. The results here are in part based upon data generated by the TCGA Research Network: https://www.cancer.gov/tcga.

## Additional Files

**Annex 1.** PanGenEU Investigators.

**Table S1.** Baseline characteristics of the study populations.

**Table S2.** Number of alleles of *HLA* Class I and Class II genes considered in the association analysis in each study population after filtering out the alleles with poor imputation quality.

**Table S3.** *HLA* allele, 95% confidence intervals bounds (ci.lb and ci.ub), odds ratios (OR), p-value, and adjusted p-value and of the association meta-analysis between the *HLA* alleles and pancreatic ductal adenocarcinoma (PDAC) risk including the PanGenEU and UK Biobank study populations.

**Table S4.** *HLA* allele, frequency (Freq), Imputation quality (INFO), odds ratios (OR), p-value, and adjusted p-value and of the association between the *HLA* alleles and pancreatic ductal adenocarcinoma (PDAC) risk in the PanGenEU study population.

**Table S5.** *HLA* allele, frequency (Freq), odds ratios (OR), p-value, and adjusted p-value and of the association meta-analysis between the *HLA* alleles and pancreatic ductal adenocarcinoma (PDAC) risk in the UK Biobank study population.

**Table S6.** Frequency in both populations (freq), Odd ratio (OR), confidence interval (CI), and p-value (p) of the inferred haplotypes from *MHC* I and II *HLA* genes significantly associated with PDAC risk by genome-wide association studies considering an additive mode of inheritance.

**Table S7.** Odd ratio (OR) and p-value (p) of the association between *HLA-A*\*02:01 and *KRAS* mutations across three different tumor types from TCGA data: PDAC, colorectal, and NSCLC (we excluded stomach because there was only one individual with G12V mutations).

**Table S8.** ICD9 and ICD10 codes for PDAC in UK Biobank

**Table S9.** Amino acid (AA), base pair (BP), Frequency (Freq), odds ratios (OR), p-value, and adjusted p-value and of the association between the HLA amino acids and pancreatic ductal adenocarcinoma (PDAC) risk in the PanGenEU study populations.

**Table S10.** *KRAS* mutational status of PDAC, colorectal, stomach, and NSCLC cancer types from the TGCA study.

**Figure S1.** Whole landscape of the alleles of *HLA* class I and II genes at two-fields in PanGenEU (Figure S1A) and in UKBiobank (Figure S1B) study populations. The x-axis shows each of the imputed *HLA* alleles across the 8 studied loci. The y-axis, represents the frequency in the PanGenEU and UK Biobank populations for each *HLA* allele.

**Figure S2.** Scatter plots of the HLA allele frequencies in PanGenEU study population versus in T1DGC, which was used as a reference panel to impute the 1-field (Figure S2A) and 2-fields (Figure S2B) *HLA* alleles in PanGenEU.

**Figure S3.** Scatter Plot of the allele frequencies in PanGenEU and UK Biobank populations, showing a moderate agreement.

**Figure S4.** Odds ratio (OR) and 95% confidence interval (CI) of the association analyses between *HLA-A*26:01* and PDAC risk and PDAC risk/protective factors in PanGenEU population. FHPC: Family history of PC; BMI: body mass index.

**Figure S5.** Venn diagram illustrating the overlap of *HLA* Class I and II gene alleles at two fields in the PanGenEU, UKBiobank, and TCGA study populations.

**Figure S6.** Hazard ratio (HR) and 95% confidence interval (CI) of the Cox proportional-hazards regression models under a codominance MoI obtained in the meta-analysis for the allele *HLA-DPB1*04* when considering PanGenEU, UK Biobank, and TCGA.

**Figure S7.** Hazard ratio (HR) and 95% confidence interval (CI) of the Cox proportional-hazards regression models obtained under the additive MoI in the meta-analysis of three populations for the 2-field alleles *HLA-A*30:01* and *HLA-DPB1*09:01*.

**Figure S8.** Boxplot showing the significant differences in IFN-γ levels between PDAC tumors with *KRAS*^G12D^ mutations vs any of the other KRAS mutated tumors obtained through the Wilcoxon test.

**Figure S9.** Hazard ratio (HR) and 95% confidence interval (CI) of the Cox proportional-hazards regression models obtained under the additive MoI in the meta-analysis of three populations for the interaction between the *HLA-A*02:01* allele with *KRAS*^G12V^ mutations.

